# Paediatric major incident triage and the use of machine learning techniques to develop an alternative triage tool with improved performance characteristics

**DOI:** 10.1101/2021.12.10.21267587

**Authors:** S. Chernbumroong, J. Vassallo, N.S. Malik, Y. Xu, D. Keene, MD. Lyttle, J.E. Smith, G.V. Gkoutos, in collaboration with PERUKI (Paediatric Emergency Research in the UK and Ireland)

## Abstract

**Background:** Triage is a key principle in the effective management of major incidents. However, there is an increasing body of evidence demonstrating that existing paediatric methods are associated with high rates of under-triage and are not fit for purpose. The aim of this study was to derive a novel paediatric triage tool using machine learning (ML) techniques.

**Methods:** The United Kingdom Trauma Audit Research Network (TARN) database was interrogated for all paediatric patients aged under 16 years for the ten-year period 2008-2017. Patients were categorised as Priority One if they received one or more life-saving interventions from a previously defined list. Six ML algorithms were investigated for identifying patients as Priority One. Subsequently, the best performing model was chosen for further development using a risk score approach and clinically relevant modifications in order to derive a novel triage tool (LASSO M2).

Using patients with complete pre-hospital physiological data, a comparative analysis was then performed comparing this to existing pre-hospital paediatric major incident triage tools. Performance was evaluated using sensitivity, specificity, under-triage (1-sensitivity) and over-triage (1-positive predictive value).

**Results:** Complete physiological data were available for 4962 patients. The LASSO M2 model demonstrated the best performance at identifying paediatric patients in need of life-saving intervention, sensitivity 88.8% (95% CI 85.5, 91.5) and was associated with the lowest rate of under-triage, 11.2% (8.5, 14.5). In contrast, the Paediatric Triage Tape and JumpSTART both had poor sensitivity when identifying those requiring life-saving intervention (36.1% (31.8, 40.7) and 44.7% (40.2, 49.4)) respectively.

**Conclusion:** The ML derived triage tool (LASSO M2) outperforms existing methods of paediatric major incident triage at identifying patients in need of life-saving intervention. Prior to its recommendation for clinical use, further work is required to externally validate its performance and undertake a feasibility assessment in a clinical context.

**What is known about this topic:** Children are frequently involved in all types of major incidents. A key principle in their management is triage, the process of prioritising patients on the basis of their clinical acuity.

Unlike in the adult population, there are currently only a limited number of paediatric triage tools for use in a major incident, with a paucity of evidence supporting their use.

A recent comparative analysis demonstrated that the adult triage tool, the MPTT-24, outperformed all existing pre-hospital paediatric triage tools at determining the Priority One paediatric patient.

**What this study adds:** We have applied machine learning algorithms to derive a novel triage tool, the LASSO M2.

This triage tool demonstrated an absolute increase in sensitivity of 52·7% over the existing UK method of pre-hospital paediatric major incident triage, the Paediatric Triage Tape.

This study has demonstrated that utilising additional parameters out with patient physiology, can lead to a marked improvement in triage tool performance.

## Introduction

Major incidents occur worldwide on a regular basis and are defined by the need for additional resources to manage either the type, severity, or location of casualties.^1^ They can arise from a variety of causes, including transport incidents, natural disasters and terrorist attacks. Children are frequently involved in all types of major incident and reminders of this in Western Europe include two terrorist attacks with over 100 paediatric casualties.^2,3^ A key principle in the effective management of major incidents is triage, the process of prioritising patients based on their clinical severity, and need for life-saving intervention (the need for which defines the Priority One patient).^4^

Whilst several methods of paediatric major incident triage exist, an increasing body of evidence demonstrates that tools currently in use in the UK are not fit for purpose. The Paediatric Triage Tape (PTT)^5^ used in the pre-hospital setting and JumpSTART^6^ advocated for the in-hospital^7^ setting both have low sensitivity for identifying paediatric patients in need of life-saving interventions, correlating with unacceptably high rates of under-triage.^8,9^

By contrast, a recent comparative analysis using trauma registry data demonstrated that the latest version of an adult triage tool (MPTT-24)^10,11^ outperforms existing pre-hospital paediatric tools in identifying the Priority One paediatric patient.^12^ There are potential benefits of extending its use to the paediatric population and applying a single triage tool across all age ranges (adult and paediatric), which include training and human factors elements for professionals responsible for decision making in such situations. However, despite high sensitivity, the MPTT-24 was associated with low specificity and positive predictive value (PPV), resulting in high rates of over-triage, which may ultimately result in an increase in critical mortality.^13^

Prior to recommending the use of an adult triage tool for use in the paediatric population, we sought to determine whether an acceptable alternative paediatric specific triage tool could be derived. With developments in artificial intelligence demonstrating improved productivity in a variety of medical applications,^14,15^ we applied machine learning (ML) techniques to a national trauma dataset to derive a novel paediatric triage tool. The key aim was to improve performance characteristics in identifying the Priority One paediatric patient, i.e. those requiring a life-saving intervention.^16^

## Methods

The Trauma Audit Research Network (TARN) database was interrogated for all paediatric (age ≤ 16 years) trauma patients, meeting TARN inclusion criteria for the ten-year period 1 January 2008 to 31 December 2017. The nature of the TARN database, and its inclusion criteria, have previously been described and are included in supplementary figure 1.^17,18^ Only patients with complete pre-hospital physiological data were included in the triage tool development. Priority One patients were defined as those receiving one or more life-saving interventions within a specified timeframe from a previously defined list (with adaptations for paediatric fluid resuscitation in keeping with Advanced Paediatric Life Support) (supplementary table 1).^16,19^ The study framework is depicted in figure 1.

**Figure 1:**
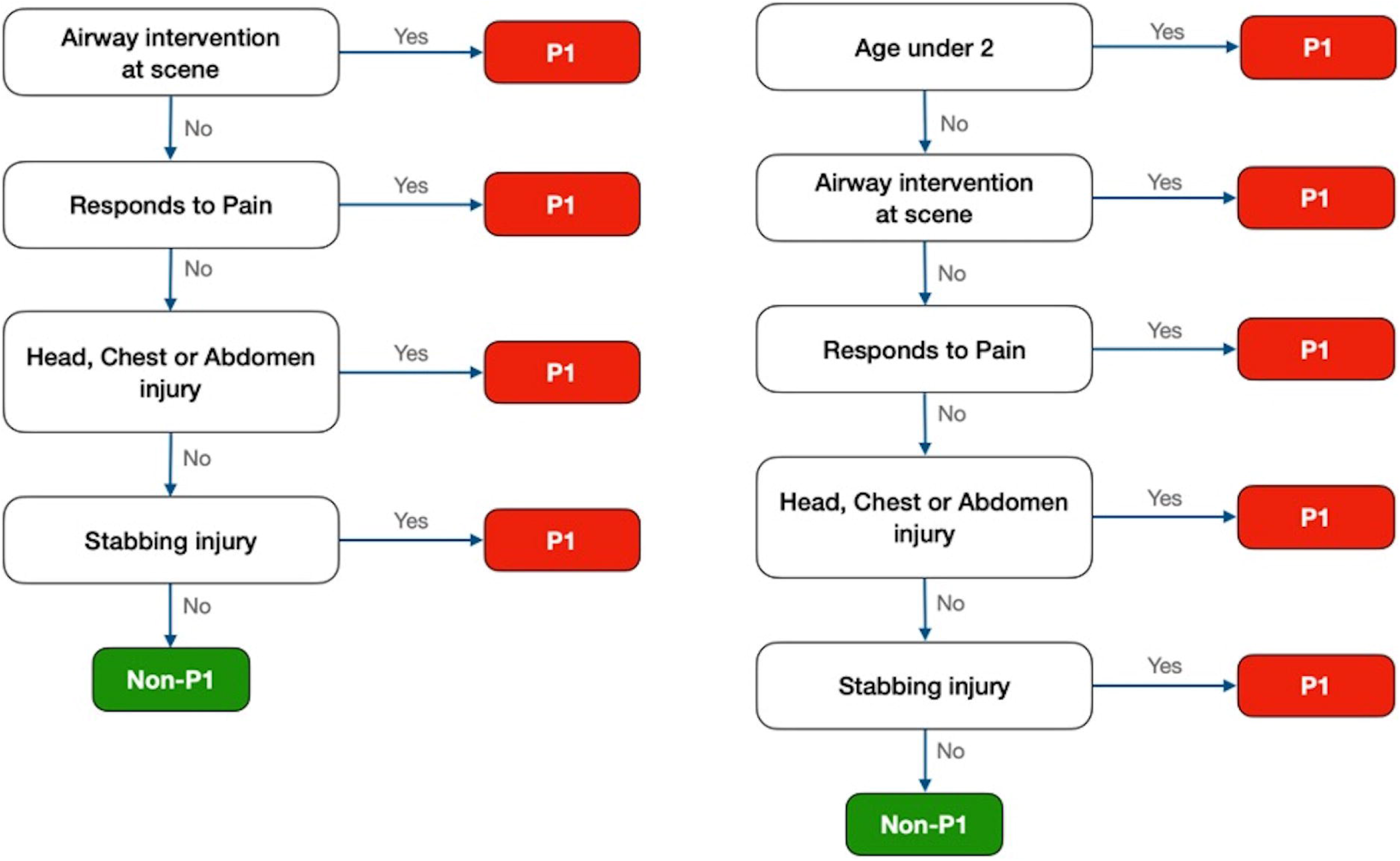
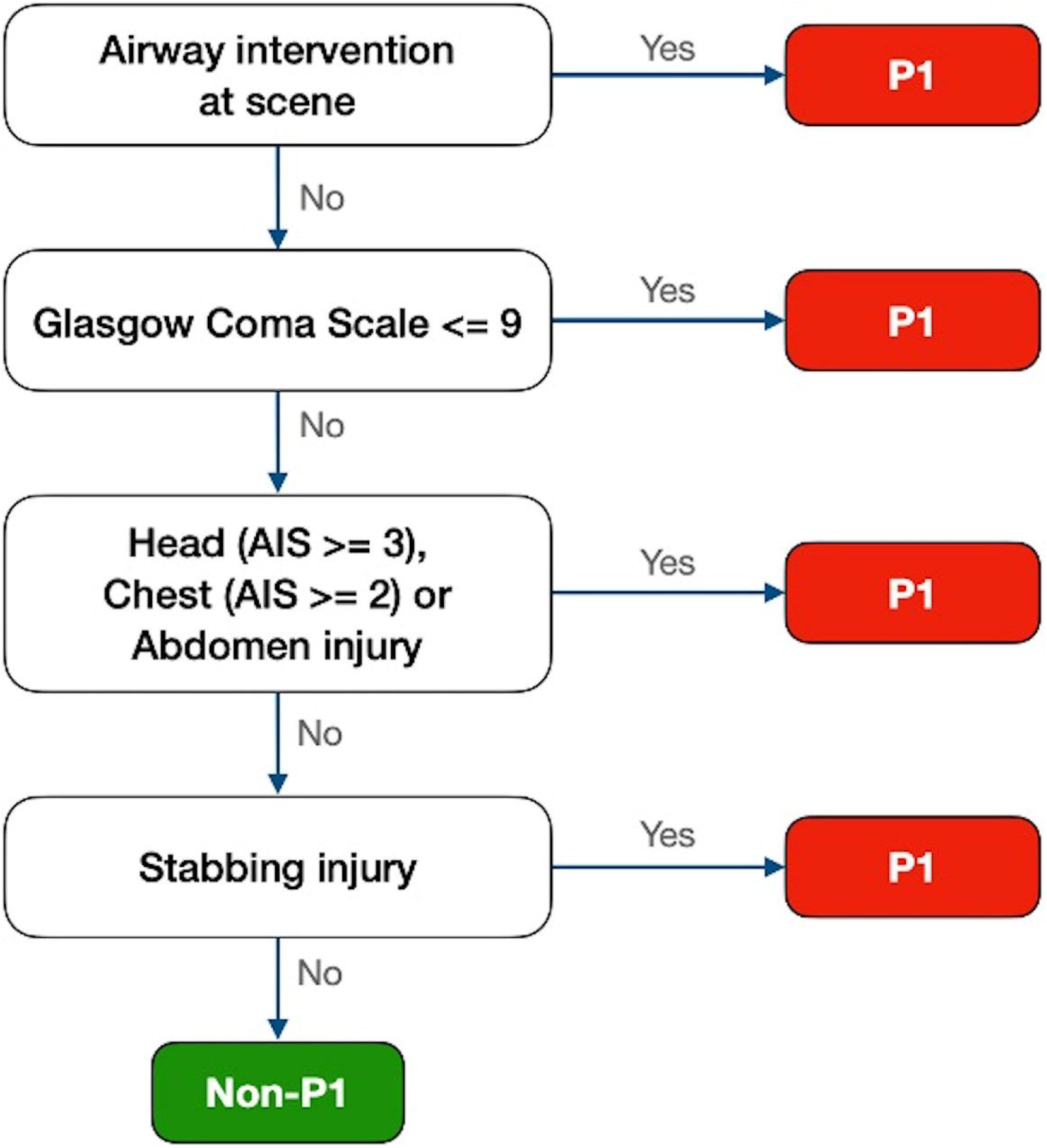
Study framework.

The dataset was subsequently split in a 70:30 ratio for training and testing respectively. Owing to an imbalance between Priority One-Not Priority One on the database, an over-sampling technique was adopted. A total of 18 parameters were used to develop ML tools (including basic physiology, age, gender, injury assessment and need for pre-hospital airway support). All hyperparameters were selected based on 5-fold cross validation. The data were normalised prior to being applied to the ML tools.

Six ML algorithms were investigated for identifying patients as Priority One: Logistic Regression,^20^ Least Absolute Shrinkage and Selection Operator (LASSO),^21^ Classification And Regression Trees (CART),^22^ Random Forest (RF),^23^ Gradient Boosting Classifier (GBC) and Naïve Bayes (NB).^24^ For tree-based algorithms, the maximum depth was set to four for the purposes of interpretability and clinical utility. An optimal cut-off was identified to maximise sensitivity and minimising over-triage. Using the testing dataset (n=1489), the performance of the ML tools was compared using area under the receiver operator curve (AUC) (figure 2 and supplementary table 2).

**Figure 2:**
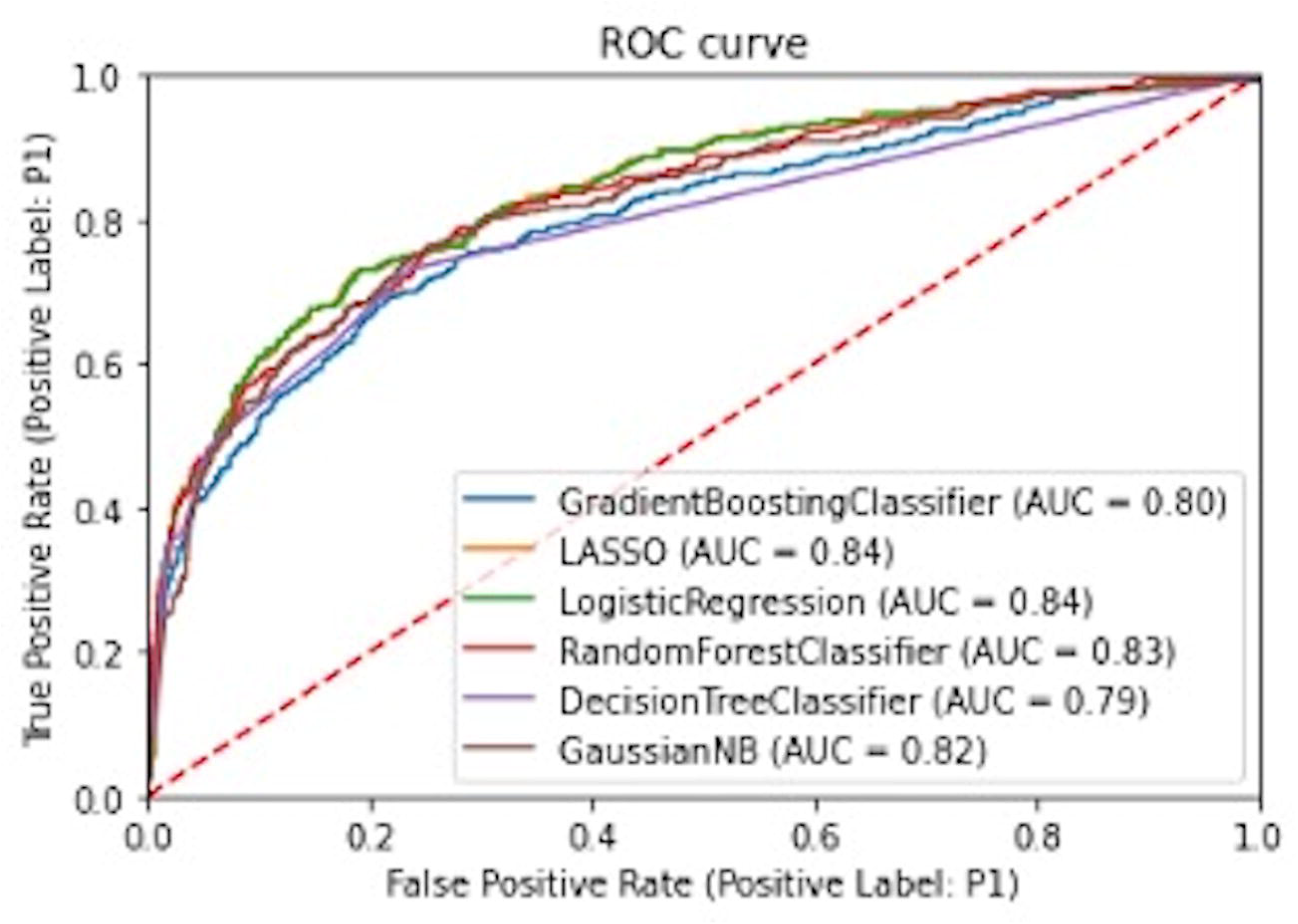
Area under curve for ML techniques.

### Primary and Secondary Outcomes

The primary outcome was correct identification of need for life-saving intervention in paediatric patients aged under 16 years in patients with complete pre-hospital physiological data. A planned secondary analysis was conducted using first recorded physiological data (pre-hospital or emergency department (ED).

In keeping with existing UK major incident doctrine^7^ where specific paediatric triage tools are utilised for those aged under 12, a subgroup analysis was conducted for this cohort of patients. An additional analysis was conducted for ML derived tools in isolation. Results from both analyses are presented as supplementary material.

### ML-driven triage tool development

Following the initial AUC analysis, owing to its high performance, simplicity, and its advantage of identifying parameters with negligible impacts on the model, the LASSO model was chosen for modification into a triage tool. Subsequently, the distribution of Priority One status across the population was explored (supplementary figure 2) and the dataset was split in two (age under four vs over four) with LASSO models derived for each using the training dataset (70% of the study population) as a basis for further tool development.

Firstly, in order to minimise the number of steps within the triage tool, only parameters with a 50% increase or decrease in odds of assigning patients as Priority One were selected for inclusion in the modified LASSO. Secondly, categorisation of GCS score (total and motor component) was undertaken based on the visualisation of percentage of Priority One patients and mortality, leading to the following categories: GCS Total 3-9, 10-12, 13-15 and GCS Motor 1-3 and 4-6 (supplementary figure 3). Thirdly, a risk score approach was applied to the modified LASSO to derive a score-based model.

Lastly, owing to the limitations of clinical triage in the pre-hospital environment during a major incident the following changes were made:

1. Replacing the GCS with the Alert, responds to Voice, responds to Pain, Unconscious (AVPU) scale.^25^
2. As it is not possible to prospectively determine the Abbreviated Injury Score (AIS)^26^ in the pre-hospital setting, injury thresholds were lowered to include any injury (AIS ≥ 1) to body regions.
3. Only parameters independently meeting the cut-off threshold were included in the derived triage tool.

These adaptations were combined to produce the triage tool LASSO M1. Owing to the distribution of Priority One status and age (supplementary figure 2), a further adaptation to LASSO M1 was made with the inclusion of a lower age threshold (two years) as an automatic trigger for Priority One status (LASSO M2). Performance of these models was then compared with existing paediatric pre-hospital major incident triage tools and the latest version of the MPTT-24 in the primary comparative analysis (table 3). A description of existing paediatric triage tools and any assumptions required for their retrospective application is provided in supplementary table 3.

### Statistical Analysis

Statistical analysis was performed to determine the sensitivity and specificity of the triage tools in detecting the outcomes of interest; under triage (1-sensitivity) and over-triage (1-positive predictive value) were subsequently calculated.^27^ 95% confidence intervals were calculated using the Wilson Score with continuity correction for binomial proportions.^28^ Python 3·7·2 was used for data processing, ML development and statistical analysis.

### Missing Data

An analysis of missing data is provided within supplementary material.

## Results

15133 patients aged under 16 met inclusion criteria, with 1197 (24·1%) receiving at least one life-saving intervention. The median age was 11·9 years (IQR 8·0-14·2), with a predominance to male gender (69·5%) and blunt trauma (95·4%), with motor vehicle collisions (49·6%) and low falls (23·9%) the leading mechanisms. Overall median ISS was 9 (IQR 9-17) with low mortality (1·1%). 4962 (32·8%) patients had complete pre-hospital physiological data recorded. Additional study characteristics are provided in table 1.

**Table 1.** Characteristics of study population and different testing cohorts.

| Variable | Category/Stats | Study Populations |  |  | Testing Datasets |  |  |
| --- | --- | --- | --- | --- | --- | --- | --- |
|  |  | Complete Population<br>(n=15133)* | Complete Pre-hospital data<br>(n=4962, 32·8%) | Complete First Available<br>Data** (n=8255, 54·5%) | Complete Population<br>n=11987 | Complete Pre-Hospital data<br>n=1816 | Complete First<br>Available data<br>n=5109 |
| **Gender** | Male | 10294 (68·0%) | 3447 (69·5%) | 5682 (68·8%) | 8093 (67·5%) | 1246 (68·6%) | 3481 (68·1%) |
|  | Female | 4839 (32·0%) | 1515 (30·5%) | 2573 (31·2%) | 3894 (32·5%) | 570 (31·4%) | 1628 (31·9%) |
| **Injury Severity Score**<br>**(median (IQR))** |  | 9 (9-16) | 9 (9-17) | 9 (9-16) | 9 (9-16) | 9 (9-17) | 9 (9-16) |
| **Age (years)**<br>**(median (IQR))** |  | 7 (2·3-12·5) | 11·9 (8-14·2) | 10·7 (5·6-13·8) | 4·6 (1·8-11·2) | 10·3 (3·9-13·6) | 8·6 (3·4-13) |
| **Outcome** | Alive | 14764 (97·6%) | 4909 (98·9%) | 8188 (99·2%) | 11649 (97·2%) | 1794 (98·8%) | 5073 (99·3%) |
|  | Dead | 369 (2·4%) | 53 (1·1%) | 67 (0·8%) | 338 (2·8%) | 22 (1·21%) | 36 (0·7%) |
| **Mode of injury** | Blunt | 14668 (96·9%) | 4733 (95·4%) | 7912 (95·8%) | 11667 (97·3%) | 1732 (95·4%) | 4911 (96·1%) |
|  | Penetrating | 465 (3·1%) | 229 (4·6%) | 343 (4·2%) | 320 (2·7%) | 84 (4·6%) | 198 (3·9%) |
| **Mechanism of injury**<br>**(n (%))** |  |  |  |  |  |  |  |
|  | Fall less than 2m | 6014 (39·7%) | 1187 (23·9%) | 2374 (28·8%) | 5309 (44·3%) | 482 (26·5%) | 1669 (32·7%) |
|  | Vehicle collision | 4721 (31·2%) | 2459 (49·6%) | 3616 (43·8%) | 3102 (25·9%) | 840 (46·3%) | 1997 (39·1%) |
|  | Fall more than 2m | 1514 (10·0%) | 645 (13·0%) | 1031 (12·5%) | 1111 (9·3%) | 242 (13·3%) | 628 (12·3%) |
|  | Blow(s) | 1270 (8·4%) | 327 (6·6%) | 601 (7·3%) | 1053 (8·8%) | 110 (6·1%) | 384 (7·5%) |
|  | Other | 1118 (7·4%) | 147 (3·0%) | 319 (3·9%) | 1044 (8·7%) | 73 (4·0%) | 245 (4·8%) |
|  | Stabbing | 229 (1·5%) | 130 (2·6%) | 196 (2·4%) | 141 (1·2%) | 42 (2·3%) | 108 (2·1%) |
|  | Crush | 160 (1·1%) | 42 (0·9%) | 73 (0·9%) | 134 (1·1%) | 16 (0·9%) | 47 (0·9%) |
|  | Burn | 44 (0·3%) | 4 (0·1%) | 12 (0·2%) | 41 (0·3%) | 1 (0·1%) | 9 (0·2%) |
|  | Shooting | 42 (0·3%) | 16 (0·3%) | 24 (0·3%) | 32 (0·3%) | 6 (0·3%) | 14 (0·3%) |
|  | Blast | 21 (0·1%) | 5 (0·1%) | 9 (0·1%) | 20 (0·2%) | 4 (0·2%) | 8 (0·2%) |
| **Priority One (n (%))** |  | 2820 (18·6%) | 1197 (24·1%) | 1813 (22·0%) | 2088 (17·4%) | 465 (25·6%) | 1081 (21·2%) |

\*Study population following application of exclusion criteria. \*\*Complete first available data refers to the presence of pre-hospital and/or first recorded hospital physiological data.

The AUC for ML models indicated comparable performance (figure 2), with logistic regression and LASSO models giving the best overall performance (AUC = 0·84 (0·82, 0·86)).

The key determinants of Priority One status from the original LASSO model were GCS Motor, GCS Total, the presence of any head, chest or abdominal injuries, a mechanism of injury that was either stabbing or a fall less than 2 metres and the need for a simple pre-hospital airway intervention (supplementary figure 4). With the dataset split by age (under-four vs over-four), the LASSO models were further developed using the variables described above. The models were trained on a 70% dataset, consisting of 3146 patients with complete pre-hospital physiological data. Performance in the over-four model was highest (AUC 0·87 vs 0·76). Despite incorporating additional variables into the under-four model performance was not significantly improved (supplementary figure 5). As a result, the over-four LASSO model was used for further tool development.

A simplified risk score was derived from the LASSO coefficients with the aim of easy calculation. An overall score of 2 was associated with high sensitivity for the need for life-saving intervention corresponding to a low rate of under triage. Within the modified LASSO model, the key determinants of Priority One status were the need for a simple pre-hospital airway intervention, stabbing as the mechanism of injury, a GCS ≤ 9 or the presence of any of the following AIS scores: Head ≥ 3, Chest ≥ 2 or Abdomen ≥ 1.

The testing dataset consisted of 1816 patients with complete pre-hospital physiological data. The subsequent LASSO model demonstrated a sensitivity and specificity of 82·2% (95% CI 78·3, 85·5) and 68·5% (66·0, 71·0) respectively, correlating with under and over-triage rates of 17·8% (14·5, 21·7) and 52·7% (49·2, 56·1). The individual components making up the LASSO model are described in full in table 2. For the purposes of practical application, a flow diagram was derived from the LASSO score-based model (figure 3).

**Table 2:** LASSO risk score model.

| Variable | Coefficient | Odds Ratio | Score |
| --- | --- | --- | --- |
| GCS Total 3-9 | 1.46 | 4.33 | 2 |
| GCS Total 10-12 | 0.00 | 1.00 | 1 |
| GCS Total 13-15 | -1.43 | 0.24 | 0 |
| GCS Motor 4-6 | 0.46 | 1.58 | 1 |
| Head AIS 3-6 | 1.21 | 3.36 | 2 |
| Chest AIS 2-6 | 0.97 | 2.63 | 2 |
| Abdomen AIS 1-6 | 0.95 | 2.59 | 2 |
| Airway intervention at scene | 1.10 | 3.00 | 2 |
| Stabbing mechanism of injury | 1.18 | 3.26 | 2 |
| Fall less than 2m | -0.73 | 0.48 | 0 |
| Penetrating mechanism of injury | 0.32 | 1.37 | 1 |

**Figure 3:**
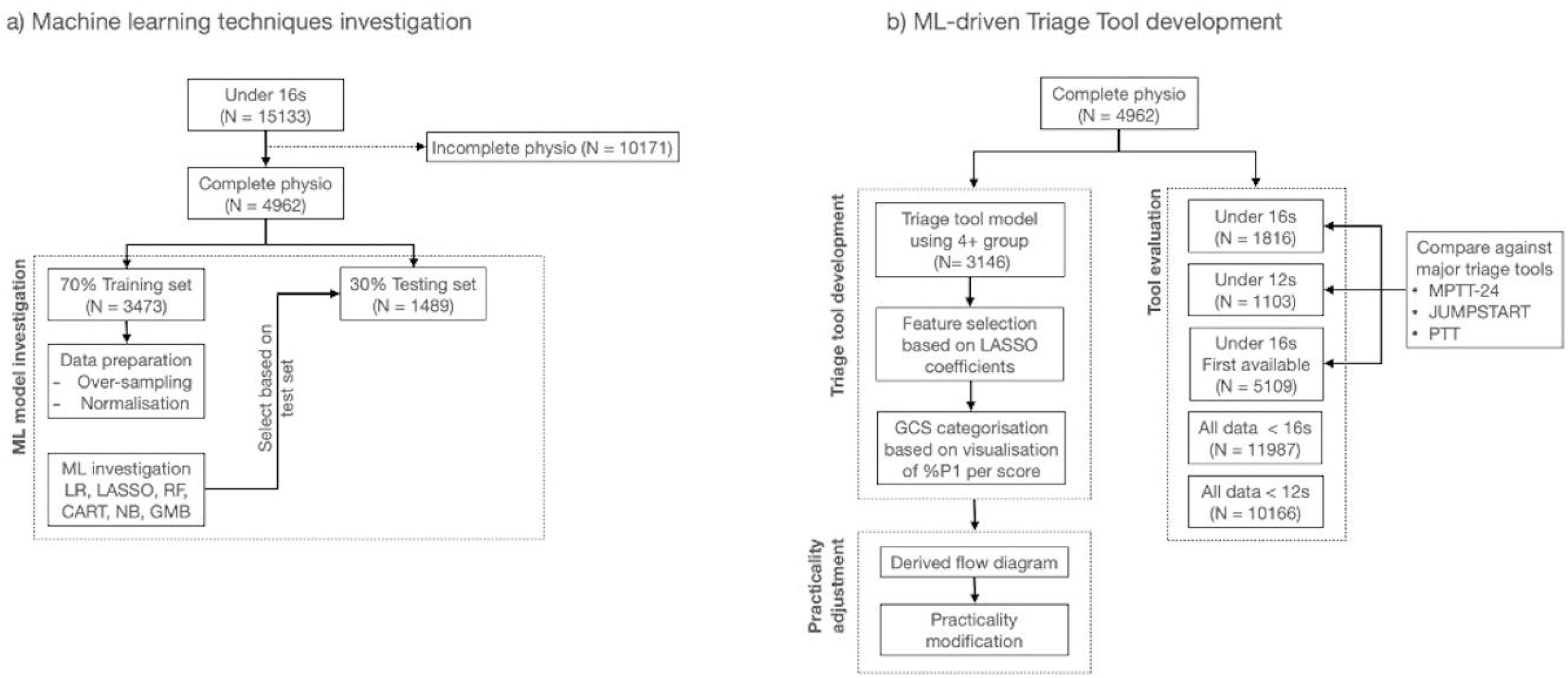
Original triage tool derived from LASSO risk score.

### Subsequent analysis following clinical review

The LASSO M1 (figure 4a) had a sensitivity of 84·1% (80·4, 87·2), correlating to an under-triage rate of 15·9% (12·8, 19·6), whilst maintaining an over-triage rate of 55·6% (52·3, 58·9). The inclusion of the lower age threshold (LASSO M2, figure 4b) resulted in an increase in sensitivity (88·8% (85·5, 91·5)) and reduction in under-triage (11·2% (8·5, 14·5)), at the expense of a slight increase in over-triage (57·5% (54·3, 60·6)).

#### Comparative analysis

4962 (32·8%) patients had complete physiological data, of which 1816 (36·6%) were included in the testing dataset used to perform a comparative analysis between the ML tools and existing triage tools. All versions of the LASSO had the greatest sensitivity, with the M2 the highest (88·8% (85·5, 91·5)), whilst maintaining a moderate over-triage rate (57·5% (54·3, 60·6)). The next best performing triage tool was the latest version of the MPTT-24 with 83·4% (79·7, 86·6) sensitivity, and over-triage rate of 68·4% (65·7, 71·0). Of all triage tools, JumpSTART had the lowest rate of over-triage (35·2% (30·0, 40·7)), but was associated with a low sensitivity (44·7% (40·2, 49·4)) and a high rate of under-triage (55·3% (50·6, 59·8)). Full performance characteristics are shown in table 3 below.

**Table 3:** Triage tool performance with 95% confidence intervals, all patients under 16 with complete pre-hospital data.

|  | Sensitivity | Specificity | Under-triage | Over-triage |
| --- | --- | --- | --- | --- |
| **MPTT-24** | 83.4 (79.7, 86.6) | 37.9 (35.3, 40.6) | 16.6 (13.4, 20.3) | 68.4 (65.7, 71.0) |
| **JUMPSTART** | 44.7 (40.2, 49.4) | 91.6 (90.0, 93.0) | 55.3 (50.6, 59.8) | 35.2 (30.0, 40.7) |
| **PTT** | 36.1 (31.8, 40.7) | 84.9 (82.9, 86.7) | 63.9 (59.3, 68.2) | 54.8 (49.6, 60.0) |
| **LASSO** | 82.2 (78.3, 85.5) | 68.5 (66.0, 71.0) | 17.8 (14.5, 21.7) | 52.7 (49.2, 56.1) |
| **LASSO_M1** | 84.1 (80.4, 87.2) | 63.7 (61.1, 66.3) | 15.9 (12.8, 19.6) | 55.6 (52.3, 58.9) |
| **LASSO_M2** | 88.8 (85.5, 91.5) | 58.7 (56.0, 61.3) | 11.2 (8.5, 14.5) | 57.5 (54.3, 60.6) |

MPTT-24 – Modified Physiological Triage Tool 24 (2018 version including airway opening manoeuvre), PTT – paediatric triage tape.

### Secondary analysis

First recorded physiological data (pre-hospital or ED) were available for 8255 patients (54·5%). Patient demographics were similar in this cohort, with males accounting for the majority (68·7%) and a median age of 10·4years (IQR 5·2-13·7). Mortality (1·1%) and ISS (median 9 (IQR9-17) were unchanged when compared to the primary analysis. The frequency of patients receiving a life-saving intervention was comparable (16·7 vs 17·6%), with advanced airway intervention again the leading intervention performed (60·1%). Additional study characteristics are provided in table 1.

The secondary analysis was conducted on a testing dataset consisting of 5109 (61·9%) patients with complete first-recorded physiological data. Triage tool performance was similar to the primary analysis, with all variants of LASSO demonstrating greatest sensitivity and lowest rates of under-triage whilst maintaining moderate rates of over-triage. Full test characteristics from the secondary analysis are provided in table 4.

**Table 4:**
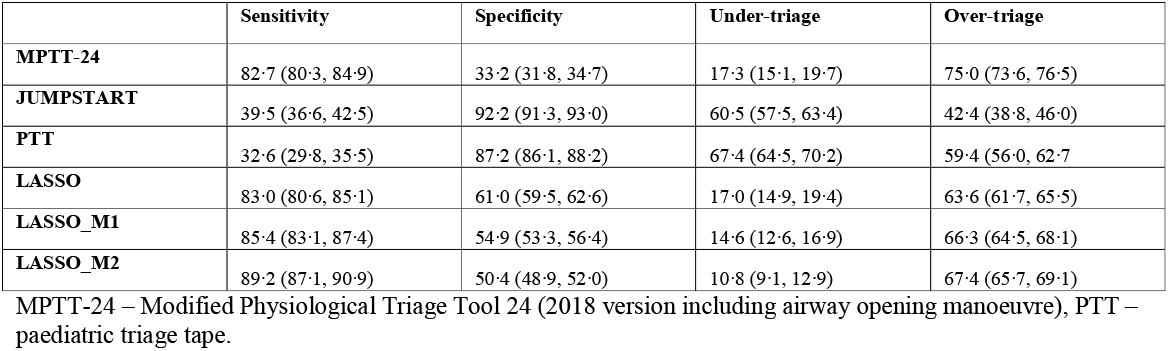
Triage tool performance with 95% confidence intervals – all patients under 16 years using first recorded physiological data.

|  | Sensitivity | Specificity | Under-triage | Over-triage |
| --- | --- | --- | --- | --- |
| **MPTT-24** | 82.7 (80.3, 84.9) | 33.2 (31.8, 34.7) | 17.3 (15.1, 19.7) | 75.0 (73.6, 76.5) |
| **JUMPSTART** | 39.5 (36.6, 42.5) | 92.2 (91.3, 93.0) | 60.5 (57.5, 63.4) | 42.4 (38.8, 46.0) |
| **PTT** | 32.6 (29.8, 35.5) | 87.2 (86.1, 88.2) | 67.4 (64.5, 70.2) | 59.4 (56.0, 62.7) |
| **LASSO** | 83.0 (80.6, 85.1) | 61.0 (59.5, 62.6) | 17.0 (14.9, 19.4) | 63.6 (61.7, 65.5) |
| **LASSO_M1** | 85.4 (83.1, 87.4) | 54.9 (53.3, 56.4) | 14.6 (12.6, 16.9) | 66.3 (64.5, 68.1) |
| **LASSO_M2** | 89.2 (87.1, 90.9) | 50.4 (48.9, 52.0) | 10.8 (9.1, 12.9) | 67.4 (65.7, 69.1) |

MPTT-24 – Modified Physiological Triage Tool 24 (2018 version including airway opening manoeuvre), PTT – paediatric triage tape.

### Sub-group analyses

#### Machine learning analysis in isolation

With the ML derived triage tools not including any individual physiological parameters (with the exception of GCS total), it was possible to analyse the performance of the ML tools on the paediatric dataset (not including those used to train the ML tools, n=11987, 79·2%). The performance of the three variants of LASSO are shown in table 5:

**Table 5:** ML-derived triage tool performance with 95% confidence intervals – all patients under 16 years.

|  | Sensitivity | Specificity | Under-triage | Over-triage |
| --- | --- | --- | --- | --- |
| **LASSO** | 84.7 (83.0, 86.2) | 68.6 (67.7, 69.5) | 15.2 (13.8, 17.0) | 63.7 (62.4, 65.1) |
| **LASSO_M1** | 86.5 (85.0, 88.0) | 62.0 (61.0, 62.9) | 13.5 (12.0, 15.0) | 67.6 (66.3, 68.8) |
| **LASSO_M2** | 90.4 (89.1, 91.6) | 49.2 (48.2, 50.2) | 9.6 (8.4, 10.9) | 72.7 (71.6, 73.7) |

The LASSO M2 continues to demonstrate the greatest performance in respect of sensitivity (90·4% (89·1, 91·6)) and the lowest rate of under-triage (9·6% (8·4, 10·9)), although this is associated with a 9·0% increase in over-triage from the original LASSO. The results from this analysis were unchanged when the dataset was constrained to patients under 12 only (supplementary table 4).

#### Comparative analysis 0-12years

The under 12 cohort accounted for 71·8% (n=10,866) of patients on the TARN database with pre-hospital physiological data available for 2516 (23·2%). The comparative analysis was performed on a testing dataset consisting of 1103 (43·8%) patients. Within this cohort, both the LASSO M2 and MPTT-24 demonstrated comparable sensitivities (88·7% (84·5, 92·0) and 88·1% (83·8, 91·4) respectively), however the LASSO M2 was associated with a lower rate of over-triage (55·9% (51·9, 59·9) vs 69·7% (66·5, 72·7)). Triage tool performance in the under 12 cohort is shown in supplementary table 5.

### Missing Data

Pre-hospital physiological data was incomplete for a significant proportion of patients (n=10171, 67.2%). Those with missing data were found to be significantly younger (median age 3·9 vs 11·9, p≤0·0001) with a higher mortality (3·1% vs 1·1%, p≤0·0001), although median ISS was unchanged between the included and excluded groups.

## Discussion

Using a large civilian trauma registry, we have used ML modelling to derive a novel triage tool that demonstrates improved performance when compared with existing paediatric major incident triage tools at identifying paediatric patients in need of life-saving interventions. The LASSO M2 method has the highest sensitivity of all triage tools, across all patient subgroups analysed, and has an over-triage rate directly comparable to that of the existing UK method of pre-hospital major incident triage. Our study findings demonstrate that ML techniques can be used to derive paediatric triage tools with improved performance over existing traditional methods in the context of a trauma registry population.

The existing literature looking at the performance of paediatric major incident triage tools is limited to either retrospective analysis of trauma registry data or prospective analysis of consecutive trauma patients.^8,9,29^ Within these settings, the results demonstrate that existing methods, such as the PTT and JumpSTART, lack the sensitivity required for clinical use; with the PTT demonstrating an absolute reduction in sensitivity of 35.8% when compared to the latest version of the adult triage tool, the MPTT-24.^8,9,12^ The results of our study here have continued to demonstrate and replicate these findings with both the PTT and JumpSTART triage methods.

The use of ML methods in an unrestrained manner allows for the identification of key parameters with which to derive a new triage tool and removes the element of author bias in selecting parameters. Within the LASSO models, except for GCS, all other physiological parameters (heart rate and respiratory rate) have been excluded as they were determined not to be predictive of need for life-saving intervention. The exclusion of these physiological assessments removes the need for physiological measurement and results in a tool reliant on the provider assessing for a response to painful stimuli and looking at the patient for anatomical injury, which will result in a more rapid triage assessment. The superior performance of ML derived tools compared with existing triage tools is likely to be attributable to the ability of ML to capture the non-linear trend within data, and additionally with the use of different cut-offs, tools can be manipulated to achieve the desired objective (e.g. high sensitivity and low rates of under-triage).

The LASSO model derived is a hybrid triage tool, incorporating all three of physiological, anatomical and mechanism of injury assessment – similar triage tools are typically used for individual field triage,^31^ but have not been advocated in the major incident context, owing to their perceived relative complexity. However, by simplifying these assessments and only including minimally invasive measurements or similar (i.e. any injury and response to pain), we have reduced the complexity of the assessment process.

The original anatomical assessment in the LASSO model, incorporated at least severe (AIS 3) or moderate (AIS 2) head and chest injuries, but as the AIS is retrospectively assigned once all investigations and procedures are complete, it lacks clinical utility and is therefore not used in the pre-hospital setting.^26^ However, by reducing the injury assessment to include any minor (AIS 1) injury, we have increased its applicability to the pre-hospital setting (LASSO M1 and M2). The practical ease of application of the LASSO M2 method is increased further by the incorporation of the AVPU scale and ‘does the patient respond to pain’ as a surrogate for GCS Total ≤= 9 included within the original LASSO model. Whilst this level is traditionally documented as occurring at a median GCS score of 8, the interquartile range is GCS 7-9, therefore for the purposes of clinical useability we deemed this to be an appropriate surrogate.^25,30^

The inclusion of a lower age threshold (LASSO M2) improves the sensitivity of the LASSO triage tool, but comes at the expense of increased over-triage, ranging from 3·8-9·0% dependent on the cohort analysed. This step was introduced owing to the increased likelihood of needing a life-saving intervention or death if aged under 2 (supplementary figure 2). We believe this is a clinically important and pragmatic step; assessment of (predominantly) non-verbal children can be difficult, is frequently emotive and this can be even more so for those with limited experience of dealing with children in this age group; all of which will undoubtedly be exaggerated in high stress situations such as a major incident. The step of automatically classifying these children as Priority One is also likely to help in reducing the cognitive burden at scene. However, we acknowledge that the introduction of this step may result in an increased rate of over-triage, although, this is likely to be negligible with the numbers of children from this cohort involved in major incidents is typically low.^2,3^

## Limitations

A key limitation of our study is the use of a retrospective trauma database with which to conduct our analyses; the leading mechanisms of injury recorded were motor vehicle collisions and low falls respectively which are unlikely to be wholly representative of that which are encountered at a major incident.^2,32^ Furthermore, owing to low numbers of patients sustaining either penetrating trauma or burns being included in the TARN database, we are unable to reliably comment on the derived tools’ performance in major incidents with a preponderance to these mechanisms, Additionally, whilst any assessment of triage tool performance should ideally be undertaken in the environment in which they are expected to function, but owing to the unpredictable nature of major incidents this is impractical. As a result, trauma databases incorporating large numbers of patients injured in isolation are frequently used as a surrogate. However, the use of trauma databases themselves are associated with limitations which, in the context of this study includes the fulfilment of inclusion criteria to be included onto the TARN database (supplementary figure 1). Although only a minority (18·6%) of patients included on the TARN database received a life-saving intervention, the presence of inclusion criteria is likely to skew the study population towards those with more severe injuries. However, we also recognise that there may be a number of children who received life-saving interventions following injury, but who did not meet the inclusion criteria of a minimum 72-hour hospital admission.

Lastly, we acknowledge that the proportion of missing data (specifically pre-hospital data) is an additional limitation of our study, with only 32·8% patients having complete pre-hospital data with which to perform our primary analysis. Whilst the median ISS was comparable between the included and excluded groups, a difference was observed in both age and mortality; those excluded were younger with increased mortality. The impact of missing pre-hospital data was mitigated through an additional analysis using first-recorded physiological data (combination of pre-hospital and ED data) where triage tool performance was unchanged. Further work is underway to perform a comparative validation using an external paediatric dataset with which to verify our results.

## Conclusion

Using trauma registry data we have demonstrated that a triage tool derived using ML techniques and incorporating simple and pragmatic clinical revisions (LASSO M2), outperforms existing pre-hospital triage tools for identifying paediatric patients in need of life-saving intervention whilst maintaining an acceptable rate of over-triage and remains simple to use. Before the LASSO M2 can be recommended for clinical use, further work is required to provide not only an external validation of its performance in an alternative dataset, but to conduct a feasibility assessment and user acceptability trial in clinical conditions.

## Supporting information

Strobe Statement

## Data Availability

De-identified patient data utilised for this study are proprietary to the Trauma Audit and Research Network, University of Manchester and may be requested directly from TARN

## Statements

### Ethical Approval

TARN has ethical approval for research on anonymised patient data. This research is governed by the code of practice approved by the Confidentiality Advisory Group appointed by the NHS Health Research Authority. TARN has approval from the Confidentiality Advisory Group under Section 251 of the NHS Act 2006. Additional individual ethical approval was not required for this research.

### Clinical Trial Registration

Not applicable.

## Acknowledgements

The authors thank Prof Fiona Lecky (research director) and Antoinette Edwards (Chief Executive Officer) at Trauma Audit Research Network (TARN) for facilitating access to the TARN database.

## Author contributions

JV and SC designed the study. JV, SC, NM, YX verified the underlying data. JV and SC conducted the analysis and interpretation. JV wrote the initial draft of the manuscript. All authors contributed with critical revisions to the manuscript and approved the final manuscript. JV takes responsibility for the manuscript as a whole.

## Patient and public involvement

Patients and/or the public were not involved in the design, or conduct, or reporting or dissemination plans of this research.

## References

1. Advanced Life Support Group. Major incident medical management and support: the practical approach at the scene. London: BMJ Books; 2011.

2. Dark P, Smith M, Ziman H, Carley S, Lecky F, Manchester Academic Health Science Centre (MAHSC) Collaborators. Healthcare system impacts of the 2017 Manchester Arena bombing: evidence from a national trauma registry patient case series and hospital performance data. Emerg Med J. 2021 Apr 22:emermed-2019-208575. doi: 10.1136/emermed-2019-208575. Epub ahead of print. PMID: 33888513.

3. Sollid SJ, Rimstad R, Rehn M, Nakstad AR, Tomlinson A-E, Strand T, et al. Oslo government district bombing and Utøya island shooting July 22, 2011: the immediate prehospital emergency medical service response. Scand J Trauma Resusc Emerg Med. 2012 Jan 26;20(1):3.

4. Wallis L, Carley S, Hodgetts CT. A procedure based alternative to the injury severity score for major incident triage of children: results of a Delphi consensus process. Emerg Med J. 2006 Apr;23(4):291–5

5. Hodgetts TJ, Hall J. Maconochie I, Smart C. Paediatric triage tape. Prehospital Immediate Care. 1998;2:155–159

6. Romig LE. Pediatric triage. A system to JumpSTART your triage of young patients at MCIs. JEMS; 2002 Jul;27(7):52–8, 60–3.

7. NHS England. Clinical guidelines for major incidents and mass casualty events. Version 2 September 2020. Available at: https://www.england.nhs.uk/wp-content/uploads/2018/12/B0128-clinical-guidelines-for-use-in-a-major-incident-v2-2020.pdf. (Accessed 2 Aug 2021).

8. Price CL, Brace-McDonnell SJ, Stallard N, Bleetman A, Maconochie I, Perkins GD. Performance characteristics of five triage tools for major incidents involving traumatic injuries to children. Injury. 2016;47(5):988–92.

9. Wallis LA, Carley S. Comparison of paediatric major incident primary triage tools. Emerg Med J. 2006 Jun;23(6):475–8.

10. NHS England Emergency Preparedness Resilience Response EPRR Clinical Reference Group 2019. ‘Triage’. Minutes of NHS England 5 June 2019. NHS England. Leeds.

11. Vassallo J, Smith JE, Wallis LA. Major incident triage and the implementation of a new triage tool, the MPTT-24. J R Army Med Corps. 2018;164(2):103–6.

12. Vassallo J, Chernbumroong S, Malik N, Xu Y, Keene D, Gkoutos GV, et al. Comparative Analysis of Major Incident Triage Tools in Children – a UK population-based analysis. Emerg Med J. Published Online First: 27 October 2021. Doi://10.1136/emermed-2021-211706

13. Frykberg ER. Triage: principles and practice. Scand J Surg. 2005;94(4):272–8.

14. Lin D, Xiong J, Liu C, Zhao L, Li Z, Yu S, et al. Application of Comprehensive Artificial intelligence Retinal Expert (CARE) system: a national real-world evidence study. Lancet Digit Health. 2021; 3(8):e486–95.

15. Bates DW, Levine D, Syrowatka A, Kuznetsova M, Craig KJT, Rui A, et al. The potential of artificial intelligence to improve patient safety: a scoping review. NPJ Digit Med. 2021;4(1):54–8.

16. Lerner EB, McKee CH, Cady CE, Colella MR, Liu JM, Schwartz R, et al. A consensus-based gold standard for the evaluation of mass casualty triage systems. Prehosp Emerg Care. 2015;19(2):267–71.

17. Kehoe A, Smith JE, Bouamra O, Edwards A, Yates D, Lecky F. Older patients with traumatic brain injury present with a higher GCS score than younger patients for a given severity of injury. Emerg Med J. 2016;33(6):381–5.

18. Barnard E, Morrison JJ, Madureira RM. Resuscitative endovascular balloon occlusion of the aorta (REBOA): a population based gap analysis of trauma patients in England and Wales. Emergency Med J. 2015;32(12):926–32.

19. Advanced Life Support Group. Advanced Paediatric Life Support. London: BMJ Books; 2016.

20. Dreiseitl S, Ohno-Machado L. Logistic regression and artificial neural network classification models: a methodology review. J Biomed Inform. 2002;35(5-6):352–9.

21. Roth V. The generalized LASSO. IEEE Trans Neural Netw. 2004;15(1):16–28.

22. Lawrence RL, remote AWPEA, 2001. Rule-based classification systems using classification and regression tree (CART) analysis. Photogramm. Eng. Remote Sens. 2010; 67(10):1137–1142

23. Liu Y, Wang Y, Zhang J. New Machine Learning Algorithm: Random Forest. In: Information Computing and Applications. Springer, Berlin, Heidelberg; 2012. pp. 246– 52.

24. Roth JA, Battegay M, Juchler F, Vogt JE, Widmer AF. Introduction to Machine Learning in Digital Healthcare Epidemiology. Infect Control Hosp Epidemiol. 2018 Dec;39(12):1457–62.

25. McNarry AF, Goldhill DR. Simple bedside assessment of level of consciousness: comparison of two simple assessment scales with the Glasgow Coma scale. Anaesthesia. 2004;59(1):34–7.

26. Greenspan L, McLellan BA, Greig H. Abbreviated Injury Scale and Injury Severity Score: a scoring chart. The Journal of Trauma. 1985;25(1):60–4.

27. Peng J, Xiang H. Trauma undertriage and overtriage rates: are we using the wrong formulas? Am J Emerg Med 2016;34(11):2191–2192.

28. Association EWJOTAS, 1927. Probable inference, the law of succession, and statistical inference. Taylor & Francis

29. Cross KP, Cicero MX. Head-to-head comparison of disaster triage methods in pediatric, adult, and geriatric patients. Ann Emerg Med. 2013;61(6):668–676.

30. Mackay CA, Burke DP, Burke JA, Porter KM, Bowden D, Gorman D. Association between the assessment of conscious level using the AVPU system and the Glasgow coma scale. Prehospital Immediate Care. 2000;4:17–9.

31. London Ambulance Service. London Major Trauma Triage Decision Tool v4.1 July 2020. Accessed 28 September 2021. Available at: https://www.c4ts.qmul.ac.uk/downloads/las-major-trauma-triage-decision-tool-(adults)-2020.pdf

32. Hunt P. Lessons identified from the 2017 Manchester and London terrorism incidents. Part Two: the reception and definitive care (hospital) phases. BMJ Military Health. 2020; 166(2):115–119

