## Supplementary tables and figures for "Paediatric major incident triage and the use of machine learning techniques to develop an alternative triage tool with improved performance characteristics"

**Supplementary Figure 1: TARN Inclusion Criteria**

**Supplementary Table 1: Lerner criteria defining life-saving interventions**

Limb-conserving surgery performed within 4 hours of arrival at hospital on a limb that was found to be pulseless distal to the injury prior to surgery

Neurologic, vascular, or haemorrhage-controlling surgery to the head, neck or torso performed within 4 hours of arrival to hospital.

Chest tube placed within 2 hours of arrival at hospital

Escharotomy performed on a patient with burns within 2 hours of arrival at a hospital

An advanced airway intervention (e.g. intubation, LMA, surgical airway) performed in the pre-hospital setting

IV vasopressors administered within 2 hours of arrival at hospital or within 4 hours of arrival at hospital

Patient who required EMS initiation of CPR (i.e. had a cardiac arrest) during transport, in the ED, or within 4 hours of arrival at a hospital

Arrived in the ED with uncontrolled haemorrhage

**Supplementary Table 2: Machine Learning derived triage tool performance with 95% confidence intervals (Testing dataset, n=1489)**

| **ML Tool** | **Sensitivity** | **Specificity** | **Under-triage** | **Over-triage** | **AUC** |
| --- | --- | --- | --- | --- | --- |
| LASSO_0·3 | 83·3 (78·9, 86·9) | 64·2 (61·4, 67·0) | 16·7 (13·1, 21·1) | 57·5 (53·7, 61·1) | 83·6 (81·6, 85·7) |
| LR_0·2 | 91·6 (88·2, 94·2) | 47·0 (44·1, 50·0) | 8·4 (5·8, 11·8) | 64·5 (61·4, 67·6) | 83·6 (81·5, 85·6) |
| RF_0·4 | 78·8 (74·2, 82·9) | 70·2 (67·4, 72·8) | 21·2 (17·1, 25·8) | 54·4 (50·3, 58·3) | 82·7 (80·6, 84·8) |
| CART_0·3 | 73·0 (68·0, 77·4) | 76·6 (74·0, 79·1) | 27·0 (20·9, 26·0) | 50·2 (45·8, 54·5) | 78·8 (76·5, 81·2) |
| NB_0·4 | 80·5 (75·9, 84·4) | 68·1 (65·2, 70·7) | 19·5 (15·6, 24·1) | 55·5 (51·6, 59·4) | 81·6 (79·4, 83·8) |
| GBC_0·1 | 94·7 (91·7, 96·7) | 22·5 (20·1, 25·0) | 5·3 (3·3, 8·2) | 72·0 (69·4, 74·5) | 79·5 (77·2, 81·9) |

ML – Machine Learning, AUC – Area Under Curve, LASSO - Least Absolute Shrinkage and Selection Operator, LR – Logistic Regression, RF – Random Forest, CART – Classification and Regression Trees, NB - Naïve Bayes, GBC – Gradient Boosting Classifier.

**Supplementary Figure 2: Priority One and Mortality Distribution by age**

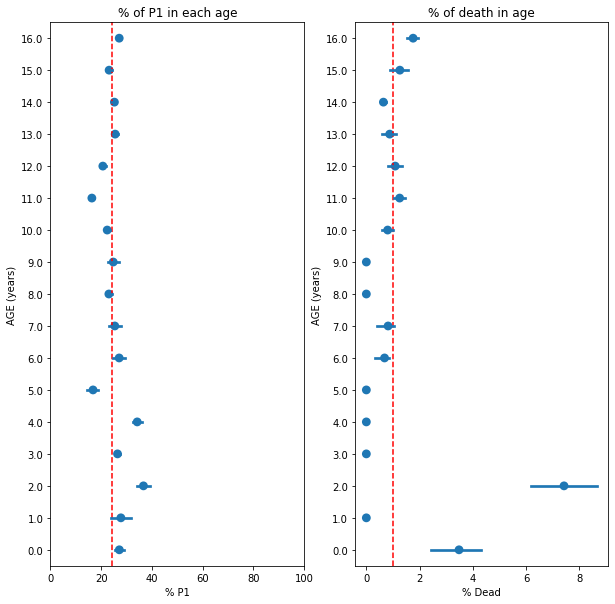

**Supplementary Figure 3: Distribution of P1 status and Mortality for GCS Motor and GCS Total**

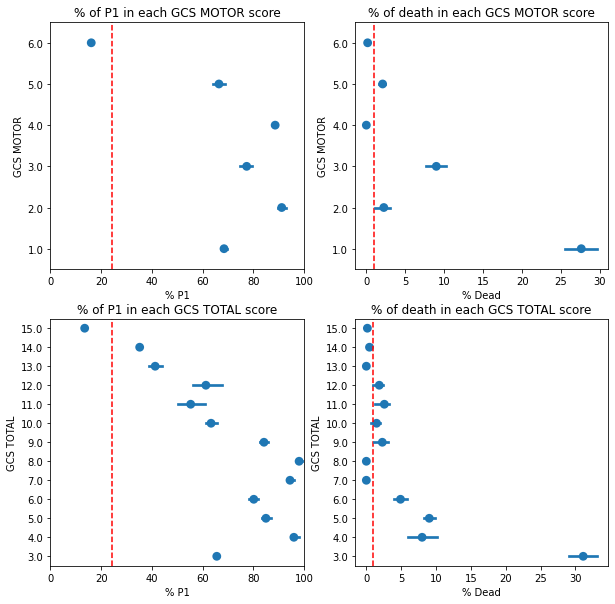

**Supplementary Table 3a: Triage Tool Comparison**

| **Tool** | **Tool components** | | | | | | |
| --- | --- | --- | --- | --- | --- | --- | --- |
|  | **1^st^ step** | **2^nd^ step** | **3^rd^ step** | **4^th^ step** | **5th step** | **6^th^ step** | **7^th^ step** |
| Paediatric Triage Tape (PTT) | <10kg:  Alert & moving all limbs  11-18kg:  Alert & moving all limbs or Walking  >19kg:  Walking | Breathing  (Open Airway if required) | Respiratory Rate:  <10kg: <20 or > 50  11-18kg: <15 or >45  >19kg: <10 or > 30 | Capillary refill < 2 sec | Heart Rate:  <10kg: <90 or > 180  11-18kg: <80 or > 160  >19kg: <70 or > 140 |  |  |
| JumpSTART | Walking? | Breathing  (Open Airway if required) | If apnoeic assess for pulse. If present give 5 rescue breaths. | Respiratory rate <15 or >45 | Palpable pulse? | Conscious level assessment (AVPU) |  |
| Modified Physiological  Triage Tool 24 (MPTT-24)* | Catastrophic Haemorrhage? | Walking? | Breathing? Open Airway if required. | Responds to voice? | Respiratory rate <12 or ≥24 | Heart rate ≥100 | - |

* The MPTT-24 was updated in 2018 following consultation with NHS England to explicitly include the ‘open airway’ step as part of the breathing assessment. This current version is currently in use in both UK military and civilian in-hospital practice (within the NHS Clinical Guidelines for Major Incidents).

**Supplementary Table 3b: Triage Tool Assumptions**

| **Conscious level** | In keeping with previously published studies, patients with a GCS ≤ 8 were categorised as unconscious, and those with a GCS < 13 were deemed to be unresponsive to voice. |
| --- | --- |
| **Obeys commands** | Patients with a GCS Motor component less than 6 were deemed to not be able to obey commands. |
| **Palpable pulse** | A systolic blood pressure of 60mmHg was used as a surrogate for the presence of a palpable pulse. |
| **Catastrophic haemorrhage** | Owing to the nature of the TARN registry, presence of catastrophic haemorrhage is not a collated variable and therefore was unable to be applied by triage tools including this assessment. |
| **Airway manoeuvre** | Patients who were recorded on TARN as having a ‘supported’ or ‘obstructed’ airway at scene were regarded as having received an airway opening manoeuvre. |
| **Weight calculations** | Where a weight calculation was required (either for classification of triage category i.e. by the PTT or for determination of fluid requirement as per Lerner criteria with APLS modification), standard APLS formula were used based on the age of the patient. |

**Supplementary Figure 4: Odds ratios for individual variables within the original LASSO model.**
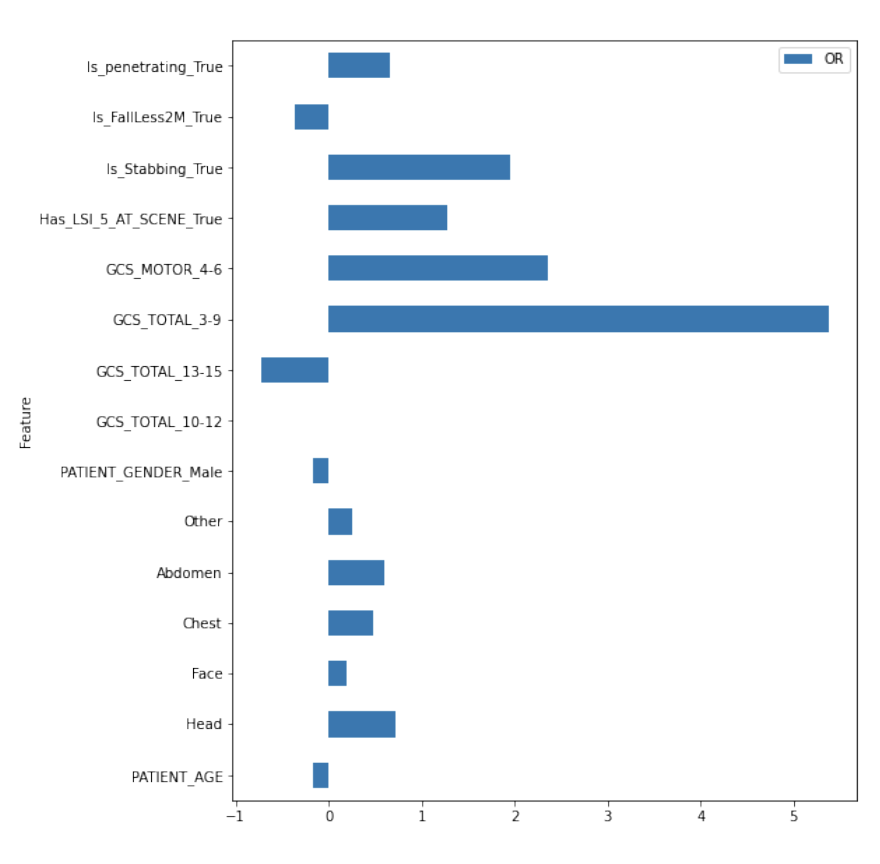

**Supplementary Figure 5: Odds ratios for individual variables within under and over-four LASSO models.**

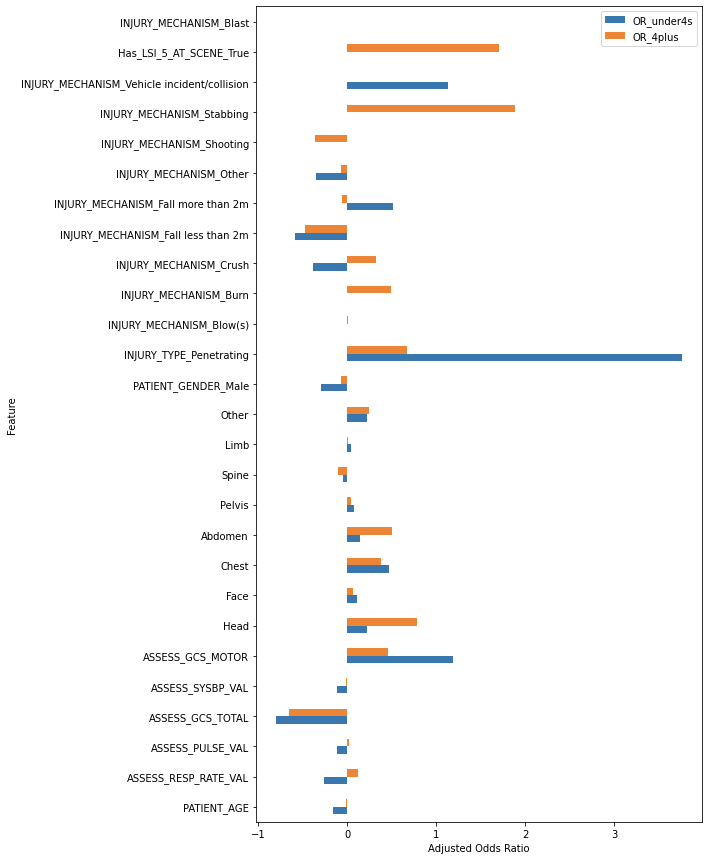

**Supplementary Table 4: Derived triage tool performance with 95% confidence intervals, - all patients under 12 – all data.**

|  | **Sensitivity** | **Specificity** | **Under-triage** | **Over-triage** |
| --- | --- | --- | --- | --- |
| **LASSO** | 83·9 (81·9, 85·7) | 71·2 (70·2, 72·2) | 16·1 | 64·6 |
| **LASSO_M1** | 86·0 (84·1, 87·7) | 63·6 (62·6, 64·7) | 14·0 | 69·2 |
| **LASSO_M2** | 91·4 (89·8, 92·7) | 47·8 (46·7, 48·9) | 8·6 | 75·2 |

**Supplementary Table 5: Triage tool test characteristics with 95% confidence intervals – all patients under 12 with complete pre-hospital data.**

|  | **Sensitivity** | **Specificity** | **Under-triage** | **Over-triage** |
| --- | --- | --- | --- | --- |
| **MPTT-24** | 88·1 (83·8, 91·4) | 23·7 (20·8, 26·9) | 11·9 (8·6, 16·2) | 69·7 (66·5, 72·7) |
| **JUMPSTART** | 41·7 (36·1, 47·5) | 91·4 (89·2, 93·2) | 58·3 (52·5, 63·9) | 35·4 (28·8, 42·6) |
| **PTT** | 35·4 (30·1, 41·1) | 82·0 (79·1, 84·6) | 64·6 (58·9, 69·9) | 57·4 (51·0, 63·5) |
| **LASSO** | 79·5 (74·4, 83·8) | 71·0 (67·7, 74·1) | 20·5 (16·2, 25·6) | 49·2 (44·6, 53·8) |
| **LASSO_M1** | 81·5 (76·5, 85·6) | 66·0 (62·6, 69·3) | 18·5 (14·4, 23·5) | 52·5 (48·1, 56·8) |
| **LASSO_M2** | 88·7 (84·5, 92·0) | 57·6 (54·0, 61·0) | 11·3 (8·0, 15·5) | 55·9 (51·9, 59·9) |

MPTT-24 – Modified Physiological Triage Tool 24 (2019 version including airway opening manoeuvre), PTT – paediatric triage tape.
